# Knowledge, risk perceptions and behaviors related to the COVID-19 pandemic in Malawi

**DOI:** 10.1101/2020.06.16.20133322

**Authors:** Jethro Banda, Albert N. Dube, Sarah Brumfield, Abena S. Amoah, Amelia C. Crampin, Georges Reniers, Stéphane Helleringer

**Author notes:** Correspondence to Stéphane Helleringer.

## Abstract

**Background:** There are limited data on knowledge and behaviors related to COVID-19 in African countries.

**Methods:** Between April 25^th^ and May 23^rd^, we contacted 793 individuals aged 18 and older, who previously participated in studies conducted in the Karonga Health and Demographic Surveillance Site in Malawi. During an interview by mobile phone, we ascertained respondents’ sources of information about COVID-19 and we evaluated their knowledge of the transmission and course/severity of COVID-19. We also asked them to evaluate their own risks of infection and severe illness. Finally, we inquired about the preventive measures they had adopted in response to the pandemic. We described patterns of knowledge and behaviors by area of residence (rural vs. urban).

**Results:** We interviewed 630 respondents (79.5% response rate). Four hundred and eighty-nine respondents resided in rural areas (77.6%) and 141 in urban areas (22.4%). Only one respondent had never heard of COVID-19. Misconceptions about the modes of transmission of SARS-CoV-2, and about the course and severity of COVID-19, were common. For example, 33.2% of respondents believed that the novel coronavirus is also waterborne and 50.6% believed that it is also bloodborne. A large percentage of respondents perceived that there was no risk, or only a small risk, that they would become infected (44.4%), but 72% of respondents expected to be severely ill if they became infected with SARS-CoV-2. Increased hand washing and avoiding crowds were the most reported strategies to prevent the spread of SARS-CoV-2. Use of face masks was more common among urban residents (22.5%) than among rural residents (5.0%).

**Conclusion:** Despite widespread access to information about the COVID-19 pandemic, gaps in knowledge about COVID-19 persist in this population. The adoption of preventive strategies remains limited, possibly due to low perceived risk of infection among a large fraction of the population.

**What is already known?:**

- SARS-CoV-2 is projected to spread widely in African countries.
- There is limited information about what affected populations know about this new health threat, and how they react to it.

**What are the new findings?:**

- In a study in Malawi, respondents lacked knowledge about several aspects of the transmission of SARS-CoV-2, and about the course and severity of COVID-19.
- These knowledge gaps were larger among residents of rural areas than among urban dwellers.
- Study respondents perceived themselves at low risk of infection with SARS-CoV-2, but they over-estimated the likely severity of the disease they would experience if they became infected.
- Most respondents reported increased frequency of handwashing, but the adoption of other protective behaviors (e.g., social distancing, use of masks) was limited, particularly in rural areas.

**What do the new findings imply?:**

- Additional information campaigns are needed to address knowledge gaps and misperceptions about SARS-CoV-2/COVID-19 in Malawi.

## INTRODUCTION

The COVID-19 pandemic has already caused more than 330,000 recorded cases and 8,000 deaths in African countries [1]. Epidemiological models have projected that close to 25% of Africa’s population could become infected with SARS-CoV-2 in 2020 [2], resulting in significant burden on health systems and between 82,000 and 189,000 deaths. As of the end of June 2020, the progression of the pandemic appears to be accelerating in several areas of the continent.

In the absence of an effective vaccine, behavioral changes are essential for limiting the diffusion and mitigating the impact of the pandemic. According to the World Health Organization [3], controlling the spread of the novel coronavirus (SARS-CoV-2) in local communities requires adopting preventive behaviors that either a) reduce the extent of contacts between population members or b) limit the likelihood that the coronavirus will be transmitted if such contact occurs. This includes, for example, maintaining an increased distance between individuals, enhancing hand hygiene or limiting mass gatherings [3]. Wearing facial masks is also increasingly recommended to limit the emission of infective droplets [4,5].

The adoption of such behaviors requires having adequate information about patterns of disease transmission and the severity of symptoms. According to standard frameworks in public health [6,7], adoption also depends on whether individuals perceive themselves as susceptible to acquiring a new disease or health threat, and whether they consider that this disease would have serious consequences for their health and well-being [8–10]. In this paper, we investigate sources of information, knowledge and risk perceptions related to the COVID-19 pandemic among a sample of Malawian adults. We then measure the prevalence of preventive behaviors during the first few weeks of the pandemic in the country.

## DATA AND METHODS

### Study setting

Malawi is a low-income country located in East Africa. Authorities declared a “state of disaster” related to the COVID-19 pandemic on March 20^th^, 2020, leading to the closure of schools and universities, for example. The country then registered its first COVID-19 case on the 1^st^ of April 2020. As of June 27^th,^ 2020, 1 152 confirmed cases had been recorded. These cases were located in all but 2 of the country’s districts. Among confirmed cases, according to the ministry of health, 632 cases were imported infections, primarily among migrant workers returning from South Africa. Four hundred and eighty-seven infections had been acquired locally [11].

We worked primarily in Karonga District, in the Northern region of the country. Karonga District is a predominantly rural district, where sources of income include fishing, farming and small-scale trading. A major highway crosses the district, leading from the Songwe border with Tanzania to Mzuzu and other large cities in the southern areas of the country. As of June 27^th^, 2020, there were 15 confirmed cases and one death in Karonga district. There were also 14 cases in neighboring Chitipa District and 99 cases in Mzuzu, the largest city of the northern region where Karonga district is located.

### Study design

The Karonga Health and Demographic Surveillance Site (KHDSS) is a data collection system that operates in the southern part of Karonga District [12,13]. Since 2002, it monitors vital events (e.g., births, deaths) that occur within a population of approximately 47 000 individuals. Prior to the COVID-19 pandemic, we have conducted several methodological studies focused on the measurement of mortality among the KHDSS population [14]. During the course of these pre-COVID-19 mortality studies, we have collected the mobile phone numbers of a number of participants to enable recruitment and/or follow-up. We used these lists of phone numbers to enroll them into a follow-up study of attitudes and behaviors towards the COVID-19 pandemic in Malawi (“COVID-19 study” thereafter). This strategy has been used in other contexts to conduct mobile phone surveys about COVID-19 attitudes and behaviors [15,16].

### Sampling

Phone numbers were available for 3 groups of respondents. First, we had the phone numbers of a sample of migrants who had left the KHDSS area. These migrants (“KHDSS migrants” thereafter) were selected at random among the lists of all former KHDSS residents who had migrated out of the area since 2002. We obtained their mobile number(s) from members of their last known household in the KHDSS area. We used these numbers to contact them so that we could confirm their interest in participating in the parent study and arrange a meeting. We then visited KHDSS migrants in their new place of residence to conduct an in-person interview in December 2019/January 2020. Second, we had phone numbers of participants in a sub-study focused on the feasibility of collecting data on mortality by mobile phone. These participants included residents of the KHDSS area (“KHDSS residents” thereafter) who were interviewed in person in February 2020. They were selected at random among residents of several population clusters located in the vicinity of Chilumba, a lakeshore town in Karonga district. Third, we asked KHDSS residents to list their maternal siblings and to refer their adult siblings to the study, so they could complete a mobile phone interview. Through this procedure, we obtained the phone numbers of a subset of the maternal siblings of these KHDSS residents. In February/March 2020, we contacted these “referred siblings” by mobile phone to conduct a short interview about health and mortality. The sample of the COVID-19 study thus includes current residents of the KHDSS area, as well as individuals dispersed throughout Malawi and residing in rural or urban areas. Individuals who had migrated outside of Malawi were not included in this study.

KHDSS residents, KHDSS migrants and referred siblings who were aged 18 years and older were eligible for the COVID-19 study. The oldest age of KHDSS residents and KHDSS migrants recruited during pre-COVID-19 mortality studies was 49 years old for women and 54 years old for men, similar to the age eligibility criteria used during Demographic and Health Surveys [17]. There was no upper limit to the age of referred siblings, however. Our sample of potential respondents in the COVID-19 study thus includes a small number of participants aged 55 and older.

Only individuals aged 18 years and older were eligible for the COVID-19 study. We sought the oral consent of respondents prior to participation in the study. This consent included a description of the goals of the study, an explanation of selection procedures and data collection procedures, and a reminder that respondents could refuse to answer any question and/or discontinue the interview at any time. The protocol for this study was approved by institutional review boards at the Malawi Ministry of Health (National Health Sciences Research Committee), at the Johns Hopkins University School of Public Health, and at the London School of Hygiene and Tropical Medicine.

### Patient and public involvement

We pre-tested study instruments with a small number of potential participants (n=15). They provided feedback on survey questions and content, which informed revisions of study instruments.

### Data collection

Due to health risks posed by in-person interviews during the COVID-19 pandemic, all data collection occurred remotely. We recruited five interviewers who had previously worked on the pre-COVID-19 mortality studies. These interviewers had extensive experience of health-related data collection. For example, they had also worked as interviewers during the 2015/16 Demographic and Health Survey in Malawi. Training for the COVID-19 study was conducted in person at the end of March, i.e., before the first confirmed COVID-19 cases were recorded in Malawi. To ensure the continuity of data collection during potential lockdowns, and to abide by local social distancing guidelines, interviewers then conducted phone interviews from their own home. Prior to the start of the study, the study supervisor visited each home to ensure that phone network coverage was sufficient and that a private space was available where interviewers could conduct interviews. Supervision procedures included daily update calls between the supervisor and each study interviewers, as well as continuous review of completed study forms. All data were collected using surveyCTO, a data collection platform widely used in population health research in low and middle-income countries. Using this platform, we pre-programmed several consistency checks and data validation routines to improve data quality.

The questionnaire covered the socio-demographic characteristics of respondents. We inquired about their recent use of healthcare services and their attendance of various places and events (e.g., funerals, churches). We then asked a series of questions about the COVID-19 pandemic, including sources of information, knowledge of transmission patterns and disease course, and preventive behaviors. These modules were adapted from instruments used in high-income countries to assess attitudes and behaviors towards the COVID-19 pandemic [18]. We also adapted several survey questions about behaviors and risk perceptions that had been asked about HIV/AIDS in prior studies in Malawi [10,19]. In listing sources of information about COVID-19, or behaviors adopted to prevent the spread of SARS-CoV-2, we let respondents spontaneously list relevant answers, without prompting them about specific sources or behaviors. Finally, the questionnaire included several modules about the survival of various relatives of the respondent [20] and the respondent’s own health. In this paper, we focus solely on data pertaining to knowledge, risk perceptions and behaviors related to the COVID-19 pandemic.

After each completed interview, we provided respondents information about the novel coronavirus, including toll-free numbers to call if they had questions about the disease or if they were experiencing symptoms. Respondents were also given 1,200 Malawian Kwachas in mobile phone credit (approximately, 1.50 US dollars) for their participation. All respondents were contacted up to 10 times by study interviewers.

### Data analysis

we described key aspects of the data collection process and data quality, including participation rates and duration of interviews. We also described the socio-demographic characteristics of study participants. This included their sex, their age group (in 10-year groupings), their economic activity in the week prior to the survey, their marital status at the time of the survey, their residence history (i.e., whether they recently moved to their current household) and their region of residence (Northern, central or southern).

The risk of exposure to COVID-19 and the availability of preventive strategies might differ between urban and rural areas. For example, in Malawi as in other low-income countries, cities might often be densely populated [21], thus making social distancing difficult to implement. In this paper, we conducted all our analyses separately by place of residence (urban vs. rural areas). Although the COVID-19 study was not designed to be representative of the rural and urban areas of Malawi, we compared the distribution of (some of) these characteristics in our study sample to the distribution of similar variables observed in a recent nationally representative dataset, i.e. the 2015/16 Demographic and Health Survey [22]. This comparison is designed to better understand the particularities of our study sample and how they might affect the knowledge, risk perceptions and behaviors reported by study respondents.

Then, we presented descriptive analyses of several variables measuring knowledge and behaviors related to the COVID-19 pandemic. First, we investigated the sources of information from which respondents had heard about COVID-19. We created a continuous variable that counts the number of information sources they reported, as well as binary variables that took a value of one if a respondent heard about COVID-19 from a specific source of information (e.g., radio) and zero otherwise.

Second, we analyzed questions that asked respondents whether they agreed with several statements about the transmission of SARS-CoV-2 (5 statements), and the course and severity of COVID-19 (6 statements). Statements about the transmission of the coronavirus included, for example, affirmations that the novel coronavirus was a respiratory virus, a waterborne virus or a bloodborne disease. Statements about the course and severity of COVID-19 included, for example, affirmations that everybody infected with the coronavirus would develop severe symptoms, or that the risk of developing severe disease was higher among the elderly. We created two variables that count the number of correct answers the respondent provided about transmission patterns and disease course.

Third, we investigated the risk perceptions of individuals. During the interview, we asked respondents how likely they believed they were to become infected with SARS-CoV-2. We asked respondents to classify this likelihood in one of four categories ranging from “no chance at all” to “almost certain”. We also asked respondents to state their individual expectations about the severity of disease, if they were to become infected themselves. We asked them to classify the expected severity across 5 categories, including “no expected symptoms”, “mild”, “moderate”, “severe” or “life-threatening”. For each category, we provided a description of associated limitations [18]. For example, we explained that a “severe” infection would likely require hospitalization, whereas individuals could go about their daily life if they experienced a “mild” infection. In our analyzes, we reported the distributions of respondents across these categories of risk assessment.

Finally, we described the behaviors that respondents reported using to prevent the spread of SARS-CoV-2/COVID-19. We created a continuous variable that counts the number of strategies reported by respondents, as well as binary variables that take a value of one if a respondent reported a specific strategy (e.g., wearing a face mask) and zero otherwise. The list of potential behaviors was adapted from a survey questionnaire used in the UK [18] earlier in the pandemic.

We compared the distributions of study variables between rural and urban residents using X^2^ tests for binary variables and t-tests or non-parametric tests for continuous variables. For statistical tests, we adjusted the calculation of standard errors to account for the clustering of observations within families. All data analysis was carried out in Stata 15.1.

## RESULTS

There were 793 individuals who were eligible for the COVID-19 study, including 230 KHDSS migrants, 106 KHDSS residents and 457 referred siblings. Between April 25^th^ and May 23^rd^, 2020, 630 of these 793 potential participants accepted to complete an interview (79.5%). The main reasons for not participating included being unable to reach the participant (n = 110) and having a wrong number (n = 23). Five potential participants refused to be interviewed, one was reported to have died, and two others had migrated outside of Malawi.

The median duration of interviews was 30 minutes (Inter-Quartile Range = 24 minutes to 38 minutes). In total, interviewers made 3,035 calls to potential respondents during the course of data collection i.e., on average 3.8 calls per potential respondent. Among those who were interviewed, the mean number of calls made was 2.6 vs. 8.4 among potential respondents who could not be interviewed.

The socio-demographic characteristics of respondents are reported in Table 1, by place of residence. Close to 60% of respondents were women. Among rural residents, 95% resided in the Northern Region of Malawi. Urban residents were dispersed throughout the country, with 31.4% and 17.9% residing in the Central and Southern regions, respectively. The percentages of respondents who resided in Karonga district were 87.3% and 17.7% for rural and urban residents, respectively. Outside of Karonga district, the districts of residence of respondents in the COVID-19 study are described in appendix A1. One in 5 respondents were young adults aged 18–24 years old, whereas <3% were adults aged 55 years and older. Respondents in rural areas were more likely to be married at the time of the interview than in urban areas (64.7% vs. 55.7%, p<0.001). Urban residents were more likely to report an economic activity outside of their household in the past 7 days (57.1% vs. 34.9%, p<0.001).

**Table 1:** characteristics of study participants.

|  | **Rural Residents**<br>**N = 490** | **Urban Residents**<br>**N = 140** | **P-value** |
| --- | --- | --- | --- |
| Gender |  |  | 0.62 |
| Men | 201 (41.0) | 60 (42.9) |  |
| Women | 289 (59.0) | 80 (57.1) |  |
| Region of residence |  |  | <0.001 |
| Northern | 467 (95.3) | 71 (50.7) |  |
| Central | 12 (2.5) | 44 (31.4) |  |
| Southern | 11 (2.2) | 25 (17.9) |  |
| Recently moved in HH? |  |  | 0.50 |
| Yes | 28 (5.7) | 6 (4.3) |  |
| No | 462 (94.3) | 134 (95.7) |  |
| Age |  |  | 0.99 |
| 18–24 | 94 (19.2) | 27 (19.3) |  |
| 25–34 | 169 (34.6) | 50 (35.7) |  |
| 35–44 | 150 (30.6) | 43 (30.7) |  |
| 45–54 | 62 (12.7) | 16 (11.4) |  |
| ≥55 | 14 (2.9) | 4 (2.9) |  |
| Marital Status |  |  | <0.001 |
| Currently married | 317 (64.7) | 78 (55.7) |  |
| Separated | 59 (12.0) | 7 (5.0) |  |
| Divorced | 25 (5.1) | 4 (2.9) |  |
| Widowed | 22 (4.5) | 4 (2.9) |  |
| Never married | 66 (13.5) | 47 (33.6) |  |
| Economic activity |  |  | <0.001 |
| Worked in past 7 days | 171 (34.9) | 80 (57.1) |  |
| Did not work in past 7 days | 319 (65.1) | 60 (42.9) |  |
| Mode of recruitment |  |  | <0.001 |
| HDSS migrants | 97 (19.8) | 73 (52.1) |  |
| HDSS residents | 86 (17.6) | -- |  |
| Referred siblings | 307 (62.6) | 67 (47.9) |  |

Notes: numbers in parentheses are column percentages
p-values are based on $\chi^2$ tests of the independence of two variables.

Compared to a nationally representative dataset, the study sample included a higher percentage of women (e.g., 59.0% vs 54.0% in rural areas, appendix A2). In rural areas, it also included a higher percentage of respondents who were separated, widowed or divorced (21.8% vs. 16.5%). On the other hand, the percentages of respondents aged 18–24 years old or 55 years and older were lower in our study dataset than in the nationally representative dataset used for comparison.

Only one respondent reported that he had never heard of COVID-19 (0.15%). Other respondents reported a median of 3 distinct sources of information about COVID-19 in rural areas vs. 4 sources in urban areas (p<0.001). In both rural and urban areas, the most common sources of COVID-related information were the radio and conversations with friends (figure 1). There were however large differences in sources of information between rural and urban residents. In urban areas, respondents reported relying more extensively on the television (64% vs. 18% in rural areas), WhatsApp groups (43% vs. 14%), Social Media (31% vs. 9%), newspapers (16.5% vs. 5%) and the Internet (18% vs. 4%). In rural areas, respondents reported obtaining COVID-related information more frequently from relatives (37% in rural areas vs. 27% in urban areas) and health facilities (37% vs. 24%). Very few respondents reported obtaining COVID-related information from the hotline(s) established by the Ministry of Health (<1%).

**Figure 1:**
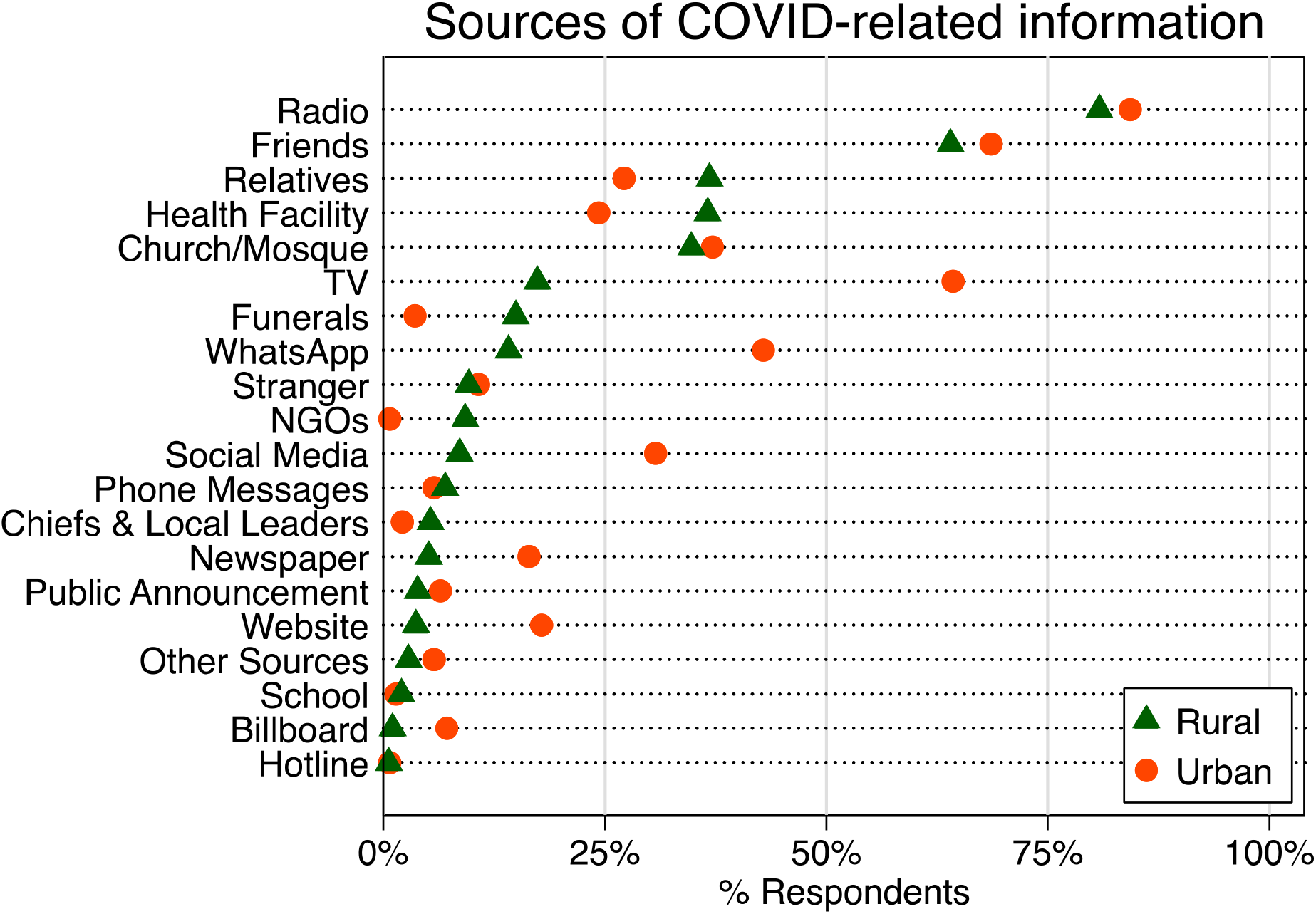
reported sources of information about COVID-19, by place of residence. *Notes:* “Phone messages” refers to informational messages sent by mobile operators (e.g., Airtel); “Hotline” refers to the toll-free numbers set up by the ministry of health to provide COVID-related information

Knowledge of transmission patterns was limited (figure 2, panel a). The median number of correct answers about transmission patterns per respondent was 3 out of 5. Close to 90% of respondents knew that the coronavirus is a respiratory virus (appendix A3), which can also be transmitted by contact with contaminated surfaces. Three out of 4 respondents knew that the coronavirus can be transmitted even if the person who is infected is not showing any symptoms. A third of respondents however believed that the coronavirus is also a bloodborne virus, whereas close to half of respondents believed that the coronavirus is also waterborne.

**Figure 2:**
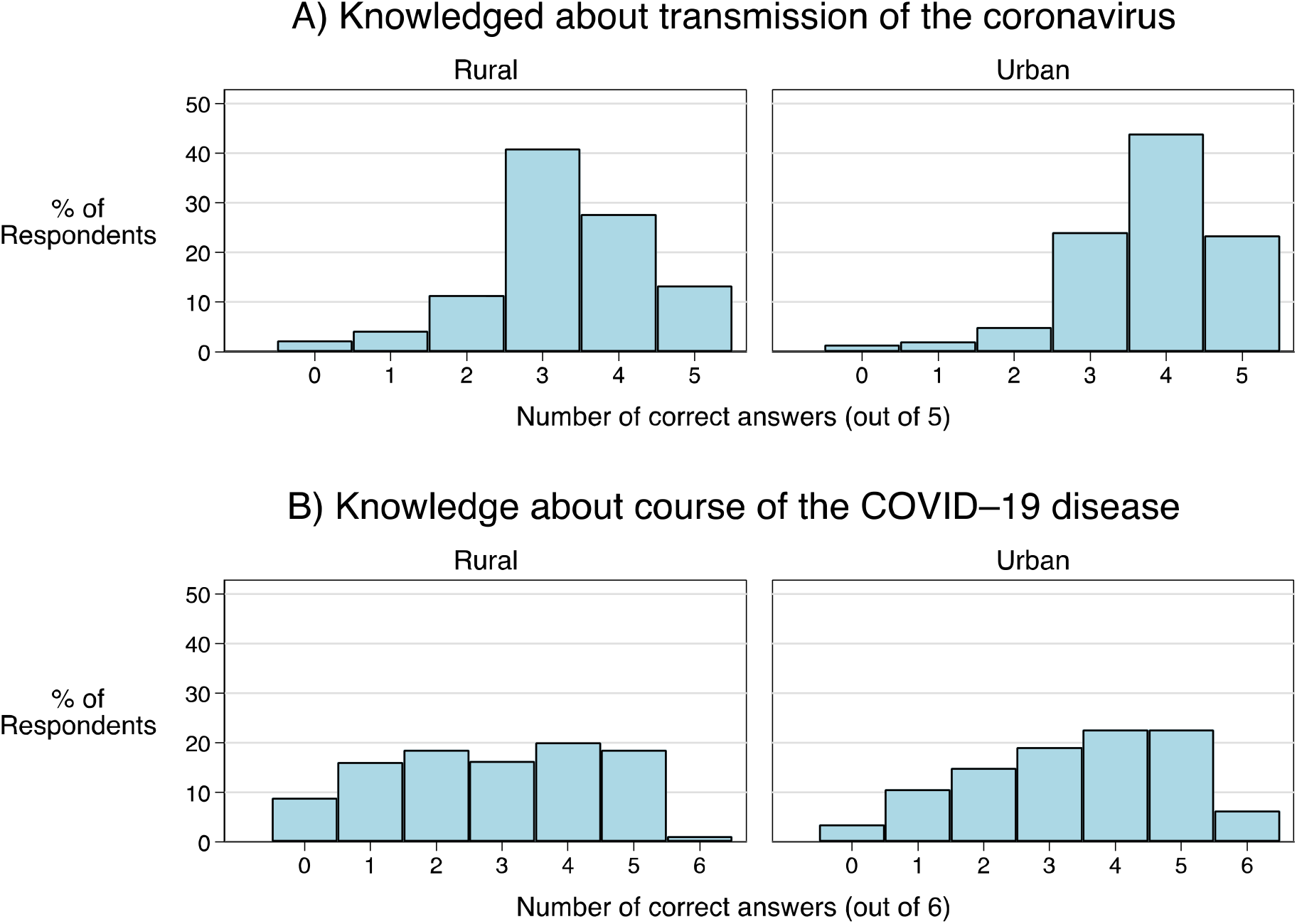
Scores summarizing knowledge of disease patterns and characteristics, by place of residence. Notes: the scores were calculated as the number of correct answers among questions concerning the transmission of the coronavirus and the course of the COVID-19 disease (see appendix A2 & A3).

Knowledge of transmission patterns was lower among respondents in rural areas (median = 3 correct answers, see figure 2) than in urban areas (median = 4 correct answers, p<0.001). The proportion of respondents who answered all questions about transmission patterns correctly was higher in urban areas than in rural areas (23.7% vs 13.3%, p<0.001). Rural respondents more frequently reported that SARS-CoV-2 is waterborne (55.2% vs. 34.3%, p<0.001; appendix A2) or bloodborne (37.0% vs. 19.3%, p<0.001).

Knowledge of the course and severity of the disease was limited (figure 2, panel b). The median number of correct answers about the course of the disease per respondent was 3 (out of 6).

One in six respondents believed that there is an effective treatment against COVID-19 and two out of three respondents believed that everyone affected by COVID-19 will ultimately become severely ill (appendix A4). Close to 40% of respondents did not know that it is possible to have and recover from COVID-19 without ever showing any symptoms. Large proportions also did not know that the risk of becoming (severely) ill varies by age: 44.8% of respondents disagreed that children under age 12 years old are less likely to become ill, whereas 32.8% of respondents disagreed that the elderly are at higher risk of severe disease.

Knowledge of the course and severity of the disease was lower among respondents in rural areas (median = 3 correct answers, see figure 2) than in urban areas (median = 4 correct answers, p<0.001). The proportion of respondents who answered all questions about course and severity correctly was lower in rural areas than in urban areas (1.2% vs. 6.4%, p<0.001). Respondents in rural areas were also less likely to know that there is no effective treatment against COVID-19 (69.5% vs. 76.4%), that SARS-CoV-2 does not always lead to severe illness (17.4% vs. 35.0%), and that people with certain chronic illnesses are more likely to become severely ill from COVID-19 (57.4% vs. 69.3%, appendix A4).

Slightly less than half of the respondents perceived themselves to be at no risk or at low risk of infection with SARS-CoV-2 (Figure 3, panel a). In rural areas, only 5.3% of respondents reported being “almost certain” to become infected, vs. 12.1% in urban areas (p=0.04). Few respondents expected to experience “no symptoms” if they ever became infected with SARS-CoV-2 (2.1%). A greater proportion of urban residents expected to experience no or only mild symptoms (Figure 3, panel b). Overall, close to 3 out of four respondents expected to experience “severe” or “life threatening” symptoms if they became ill with COVID-19.

**Figure 3:**
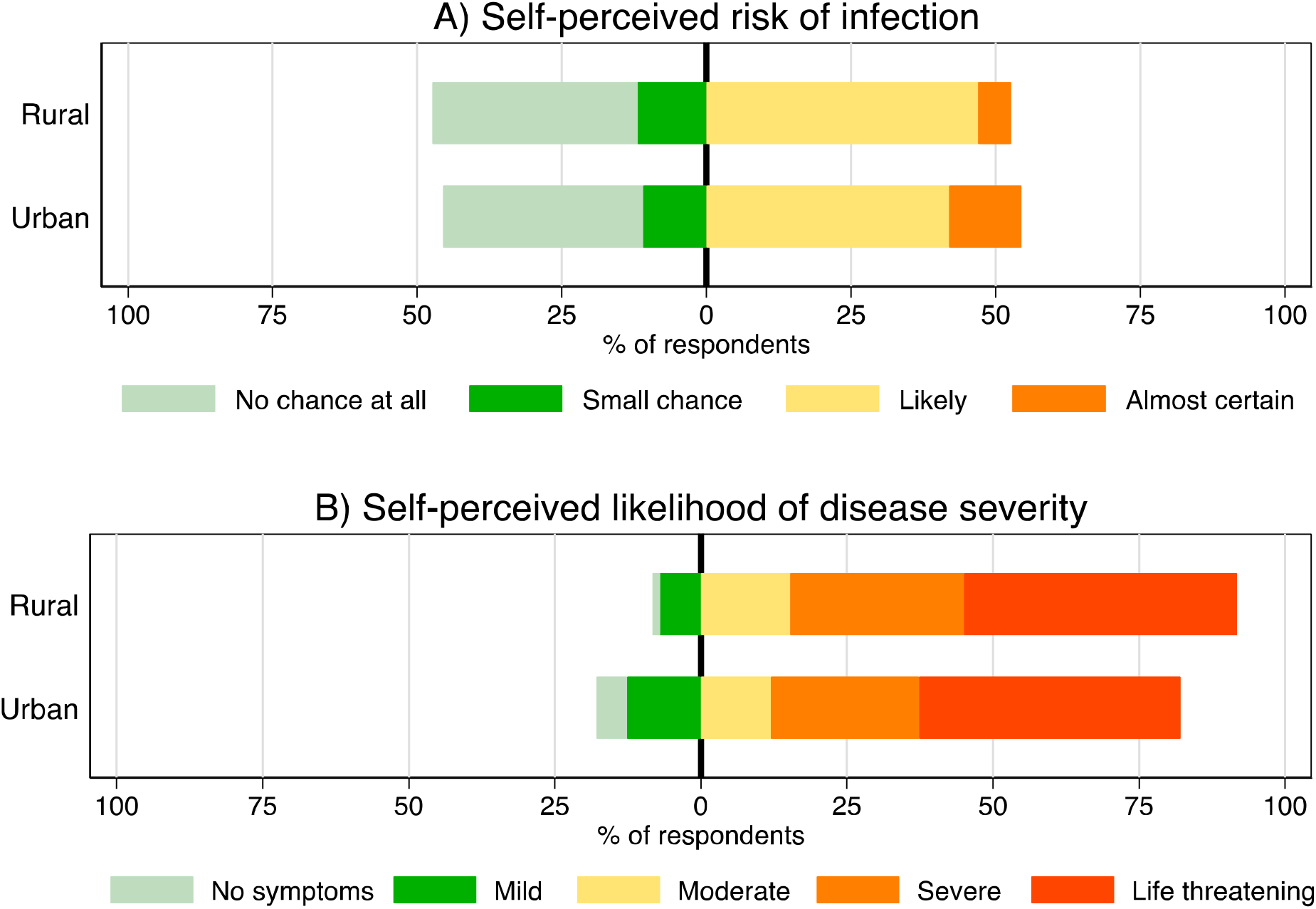
self-perceived risk of disease outcomes, by place of residence. Notes: the difference in the distribution of answers between rural and urban residents was significant the p<0.05 level according to a X^2^ test of the independence of two categorical variables for the self-perceived likelihood of disease severity. The p-value of a similar test was 0.059 for the self-perceived risk of infection.

Seven respondents reported not having adopted any strategy to prevent the spread of SARS-CoV-2 (1.1%). Among the rest of the respondents, the median number of preventive behaviors adopted was 3. Rural residents reported adopting a median number of 2 preventive behaviors, vs. 3 for urban residents (p<0.001). More than 95% of respondents reported washing their hands more frequently (Figure 4), and approximately 50% reported avoiding crowds. Only one out of five rural residents and one out of four urban residents reported staying at home. The use of face masks and hand sanitizers was more prevalent among urban residents (22.3% and 27.3%, respectively) than in rural areas (5.0% and 10.6%, respectively). Very few respondents reported avoiding hospitals and health facilities to avoid becoming infected with SARS-CoV-2.

**Figure 4:**
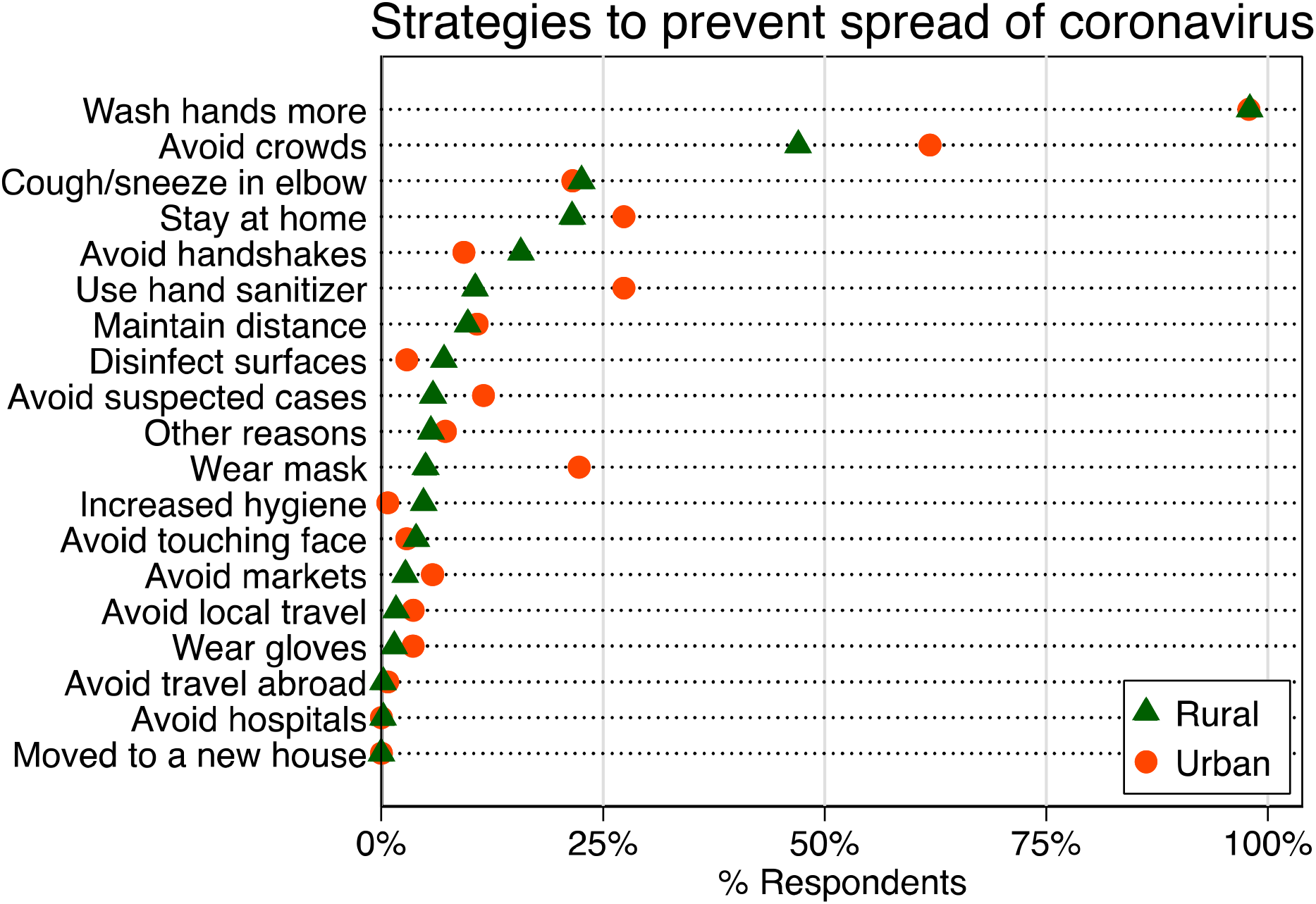
adoption of preventive strategies, by place of residence. *Notes:* percentages are calculated among respondents having adopted at least one prevention strategy (n = 622).

## DISCUSSION

Our study documented that despite widespread access to information, respondents in this Malawian population had imperfect knowledge of the patterns of SARS-CoV-2 transmission and of the course and severity of COVID-19. In particular, a large percentage of respondents believed that SARS-CoV-2 was also waterborne or bloodborne, and that COVID-19 necessarily led to severe symptoms. Knowledge of the transmission patterns and of the course/severity of COVID-19 was more limited among residents of rural areas.

Close to half of study respondents perceived themselves at no risk or at low risk of acquiring the novel coronavirus. Even in urban areas, only 1 in 8 respondents reported perceiving themselves at high risk, whereas in recent studies in Nairobi slums, this proportion was as high as 1 in 3 [15]. Compared to available epidemiological models, study respondents also seemed to over-estimate the risk of severe outcomes from COVID-19. Whereas 3 out of 4 respondents (72%) stated that they expected to experience severe or life-threatening symptoms (requiring hospitalization) if they became infected with SARS-CoV-2, a recent model of the World Health Organization [2] indicated that only 2% of the infections with SARS-CoV-2 projected to occur in Malawi in 2020 would require hospitalization. For comparison, in the United Kingdom in March 2020, only 1 in 5 respondents in a nationwide survey reported that they would expect COVID-19 to be severe or life-threatening [23], even though the population of the UK is more vulnerable to adverse disease outcomes due to its older age structure [24].

In the first few weeks of the COVID-19 pandemic in Malawi, the adoption of preventive behaviors was limited in this population. Virtually everybody reported washing hands more often, but only a limited number of respondents reported implementing other strategies including social distancing or wearing face masks. This was particularly so in rural areas, where respondents reported having adopted fewer strategies to prevent COVID-19 than their relatives in urban areas. This limited adoption of preventive behaviors might in part be related to how individuals assessed the health threat from the pandemic. Low perceived levels of infection risks have been associated with lack of behavioral changes for diseases such as influenza [8] or HIV [25,26]. Similarly, misperceptions about the likely impact of a disease on health and survival might preclude the adoption of safer practices and behaviors [27].

Our study has several limitations. First, it is based on a sample of individuals who could be reached by mobile phone. As a result, it might exclude members of the more impoverished households of the study communities. If the knowledge and behaviors of population members who do not have access to mobile phones differs from those of other members, then our study results might be biased. Second, our sampling frame was constituted from lists of participants in studies conducted in 2019/2020. Only approximately 45% of the sample was randomly selected (HDSS migrants and HDSS residents). Other respondents were referred to this study by their (randomly selected) siblings. As such, our sample is not representative of a specific population in Karonga District or elsewhere in Malawi. In particular, whereas our sample included respondents in a number of cities of the country (e.g., Lilongwe, Mzuzu), it was not representative of urban areas in Malawi. In appendix A2, however, we documented how our study sample differs from representative samples of households in rural and urban areas. The most salient difference was the limited number of respondents aged 55 years and older in our study sample. Since members of such age groups are at an increased risk of adverse COVID-19 outcomes [24], future studies of knowledge and behaviors related to the pandemic in African countries should ensure that they are adequately represented. Third, we did not investigate other determinants of the adoption of preventive behaviors according to the health belief model. For example, we did not explore the barriers and constraints that might prevent individuals from maintaining social distancing norms, from consistently using hand sanitizer or from adopting other preventive practices. This includes, for example, housing conditions, access to water sources and/or financial resources to purchase masks. Fourth, our analyses are based on self-reported data on attitudes and behaviors. These data might be affected by social desirability biases [28]. For example, respondents might over-report the number of preventive behaviors they have adopted if they believe that this might be the more acceptable answer. Sixth, we only investigated whether knowledge and behaviors varied by place of residence. We did not consider other correlates including age, marital status or economic activity. Finally, our study results might be affected by non-response during the mobile survey. Only 80% of the individuals for whom a phone number was available participated in the mobile interview. If the characteristics and behaviors of non-participants differ from those of participants, the results from the study might be biased.

Our study has important implications. It indicates that additional information campaigns are needed to address knowledge gaps about the transmission of the novel coronavirus, and the severity/course of the disease it causes. Whereas prior studies of attitudes and behaviors related to the COVID-19 pandemic have focused on urban settings [15], we also highlight that several knowledge gaps are larger, and the adoption of preventive behaviors is slower, in rural areas. Information campaigns and behavioral change communication about COVID-19 thus need to be tailored to these different contexts. For example, messages diffused using social media or WhatsApp groups are unlikely to reach a significant percentage of rural residents in Northern Malawi and similar settings. Finally, our study indicates that individual risk perceptions do not currently align with projections of the impact of the COVID-19 pandemic in Malawi: a large percentage of population members might under-estimate the risk of becoming infected, while most respondents over-estimate the risk of developing severe illness if infected. Future health communications and information campaigns related to the pandemic should address these misperceptions. According to common public health frameworks (e.g., health belief model), this might help foster the adoption of preventive behaviors in affected populations.

## Data Availability

All study data will be posted on a public repository as soon as possible.

